# Erosion of representativeness in a cohort study

**DOI:** 10.1101/2020.02.13.20022012

**Authors:** M.D. Christodoulou, J.A. Brettschneider, D. Steinsaltz

**Affiliations:** Department of Statistics, University of Oxford, Oxford, U.K.; Statistics Department, University of Warwick, Coventry, U.K.

**Keywords:** Bias, attrition, representativeness, fertility, age at menarche, age left full time education, birth cohort, 1958 British Birth Cohort, NCDS, dropouts

## Abstract

**Background:** The National Child and Development Study (1958 British Birth Cohort) follows the lives of over 17 000 people born in a single week in England, Scotland, and Wales. Since the initial recruitment there have been nine sweeps to gather subsequent life-course data, and a Biomedical Sweep (Age 44) – between Sweeps 6 (Age 42) and 7 (Age 46) – that has found widespread application in genetic studies. Due to its non-selective recruitment, the survey is frequently used as a representative proxy for the British population in demographic, epidemiological, and medical studies. We examine the effect of attrition on representativeness of female fertility and education length.

**Methods:** We compare numbers and timings of fertility-related events of female cohort members with national estimates. Spline approximation was used to link records with different aggregation intervals. Participants present in the Biomedical Sweep (Age 44) were compared to those who were not.

**Results:** We established that both timings and counts of maternities and terminations in the cohort diverge from the patterns of their contemporaries. For women who participated in the Biomedical Sweep (Age 44), we noted positive correlations of study continuation with years spent in full time education, and with age at first birth. We determined that women who did not participate in the Biomedical Sweep (Age 44) reported different fertility patterns from those who did.

**Conclusions:** It is possible to use National Statistics to quantify various forms of selection bias that inevitably creep into even the most meticulously sampled longitudinal study, and the misreporting that affects particular questions. While the 1958 Birth Cohort has reasonably been described as “broadly representative” of the British population, such a characterisation becomes questionable when the data are to be applied to questions about female fertility.

**KEY MESSAGES:**

- Fertility patterns of women reported in later sweeps of the 1958 British Birth Cohort diverge from those of their contemporaries as estimated from national statistics using spline approximation. Both maternities and terminations are underreported.
- Female participants who dropped out earlier follow national maternity trends more closely than those who participated in the study longer, but they still report slightly fewer maternities than national statistics suggest.
- Female participants who persisted through later sweeps experienced first births later and left education at a later age than those who dropped out earlier.
- Although the 1958 British Birth Cohort has been judged as representative of the British population for some research questions, the cohort population presents increasingly biased fertility patterns in female participants over time. Studies related to fertility using data from this cohort may require adjustment.

## BACKGROUND

The 1958 British Birth Cohort (National Child Development Study) was originally designed as a one-off study focusing on factors surrounding perinatal mortality in Great Britain, and including all births occurring in England, Scotland, and Wales during the first week of March 1958 (1). Cohort members have been followed throughout their lives, creating a rich dataset with extensive records on multiple aspects of their lives, such as health, education, and employment. The non-selective inclusion of every child born in Great Britain in one week has made it an attractive starting point for longitudinal studies of a representative cross-section of the British population. A large and diverse collection of variables has been amassed for the 17 415 initial cohort members (and the 925 added in the replenishment sweeps at ages 7, 11, and 16 for children immigrating to Great Britain), documenting every facet of their lives, from gestation through late middle age. Despite dropouts (due to mortality, emigration, or other reasons), more than half of the cohort (9 377 members) persisted to the Biomedical Sweep (age 44), and more than 40% participated in the most recent sweep in 2013. Levels of attrition are illustrated in Figure 1.

**Figure 1:**
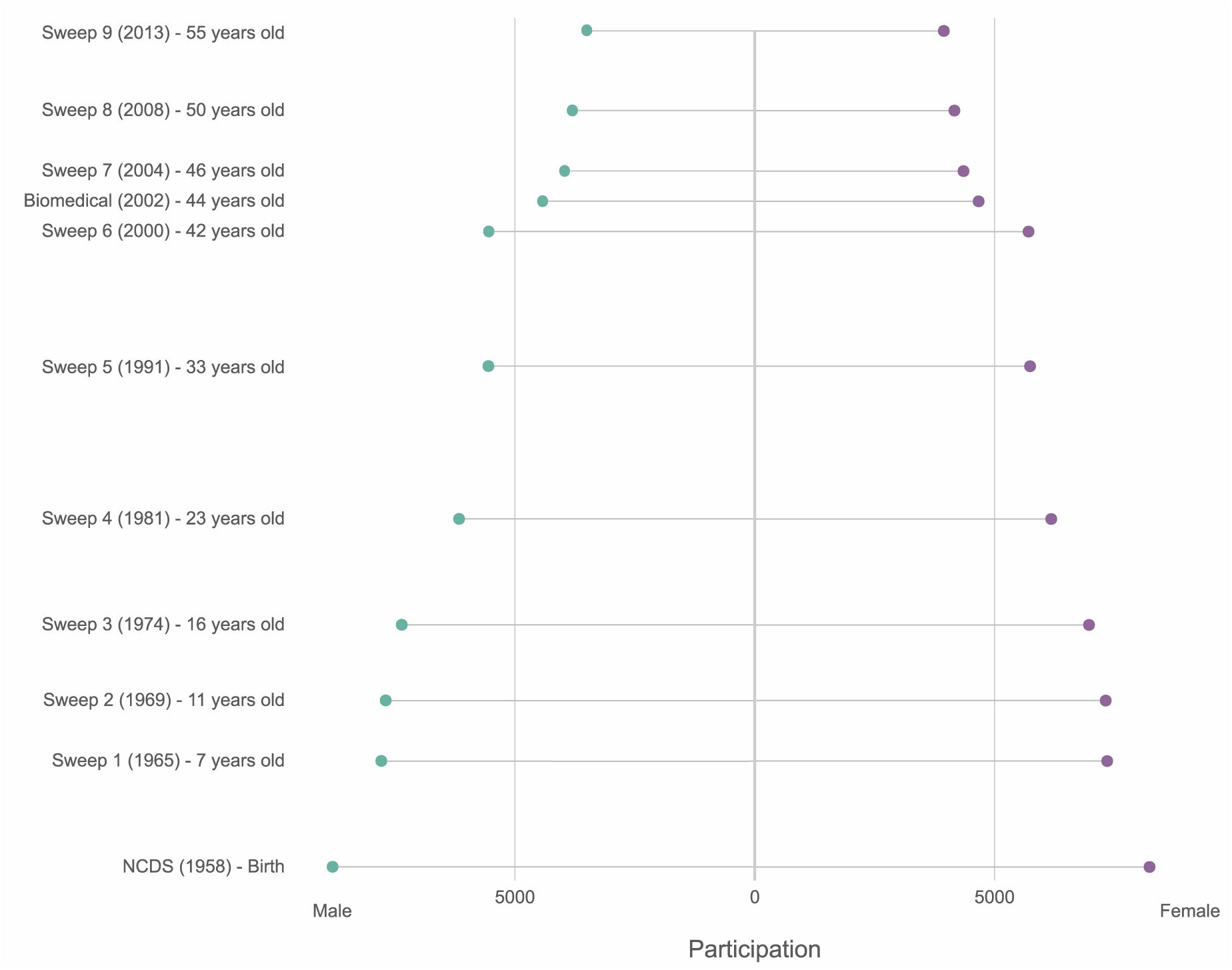
Male and female productive participation by sweep for the 1958 British Birth Cohort (National Child Development Study – NCDS). Starting with 17 415 cohort members at recruitment (8 411 female productive participants and 9 004 male), productive participation declined to 9 377 members for the Biomedical Sweep (4 712 female participants and 4 665 male) in 2002. Age of cohort member and year of Sweep are listed as the start year for each sweep. The highest average annual rate of loss occurred between Sweep 6 and the Biomedical Sweep. This pattern is consistent for both female and male participants. Productive participants are the cohort members who were successfully contacted and responded to a Sweep’s questions. As only female participants were released for this study, the male participation was calculated using the publicly available achieved samples for the cohort. The lengths of the lines are proportional to the numbers of individuals they represent.

Because of the outsized role that this cohort has played in a wide range of research, the development of its membership over time has received considerable attention. Atherton et al. (2) compared a variety of sociodemographic, health, and behavioural variables between Sweep 1 (Age 7) and the Biomedical Sweep (Age 44). They concluded that subject attrition had not substantially vitiated the broadly representative composition of the persisting cohort. This claim has been echoed in numerous publications referring to the cohort as “nationally representative” (3–9) or “broadly representative”, subject to attrition (10–16).

The present work examines whether specific data biases may distort the picture of particular research questions within an otherwise representative sample. Our focus is on the impact of attrition on fundamental indicators of education and fertility, such as age at menarche and years spent in education. We explore associations between these variables and the length of continuing participation in the study, for women present in the Biomedical Sweep (Age 44). We further compare national statistical records to the maternity and termination reports of female participants, separately for those who persisted until the Biomedical Sweep (Age 44) and those who did not. Our objective for this is to establish how the two groups compare to one another, and whether they diverge in parallel from national statistics, or follow predictable patterns. As the Biomedical Sweep (Age 44) was focused on collecting biological material and clinical data, it is only participants in this Sweep whose information is routinely released for research that aims to include genetic data. The genetic data commonly used were initially generated by the Wellcome Trust Case Control Consortium (WTCCC) studies (17,18) (Figure 2), which used members of the 1958 Birth Cohort as part of the control group for large meta-analyses on common diseases. These were subsequently made available to other researchers, and have since been used as part of control groups (usually in combination with UK Blood Donors) for a variety of medical studies, including work on leukaemia (19,20), schizophrenia and rheumatoid arthritis (22), as well as for sociological and demographic research (23,24). As these studies include only participants in the Biomedical Sweep (Age 44), we take as our primary cutoff. Our crucial aim is to better understand the composition of this group, that has served as a control in so many medical studies, and how it compares to the remainder of the cohort.

**Figure 2:**
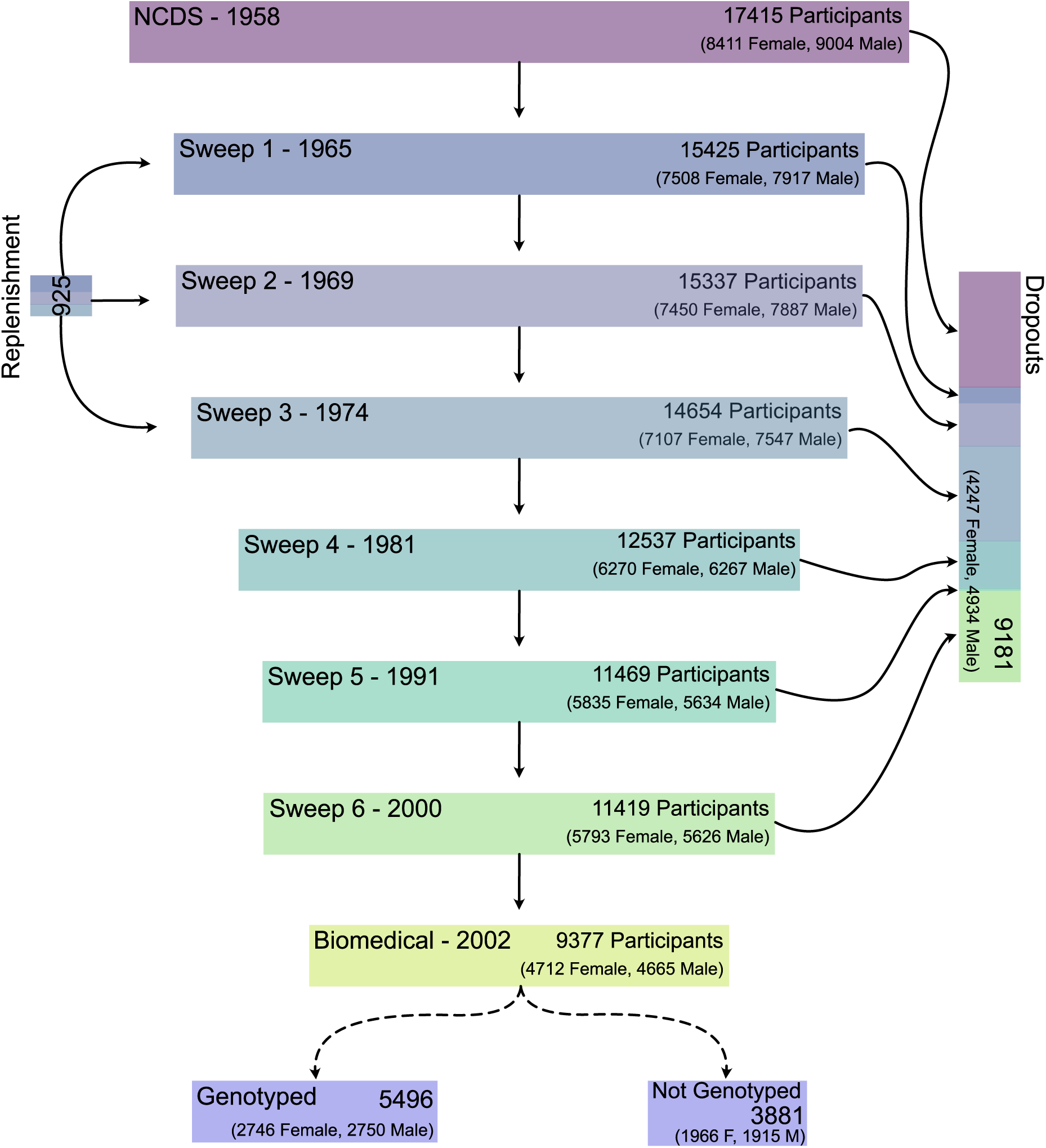
Male and female productive participation, dropouts, and replenishment in the 1958 British Birth Cohort until the Biomedical Sweep, culminating in the numbers of individuals that were genotyped. Productive participants are the cohort members who were successfully contacted and responded to a sweep’s questions. As only female participants were released for this study, the male participation was calculated using the publicly available achieved samples for the cohort. Lengths of rectangles are proportional to the total number of individuals participating.

Whether representativeness as a concept matters for epidemiological, sociological, or demographic research has been extensively debated (25–30), and is beyond the scope of this work. We are not questioning whether overall the 1958 British Birth Cohort is a representative proxy of British population for its age group. Nor are we questioning the appropriateness of the cohort members specifically as a control group for the genetic studies designed by WTCCC. Here we consider the essential yet sometimes neglected distinction between a population being representative overall, versus being representative for a particular research objective. In this work we achieve two aims. First, we examine the representativeness of the cohort in matters of female fertility, which is interesting on its own merit as well as a fundamental vital statistic that can be verified against National Registry Data. We use education as a sociodemographic variable, as a plethora of studies have demonstrated the links between education and fertility behaviour(31–34). Second, we model how researchers can benchmark a study against available national statistics in order to quantify the divergence between the studied group and their contemporaries.

We have instrumentalised the general question of representativeness in two specific questions:

1. whether annual rates of maternity and termination reported by participants who persisted through the later sweeps diverged from rates reported by those who dropped out earlier, and how each of these comports with national statistics. Due to different age aggregation in the latter, we used spline approximation to estimate annual rates, described in the Section: “Annual maternity and termination rate estimates”; and
2. whether participant attrition interacts with education and fertility variables. This was conducted using time-to-event analysis, and is presented in the Section: “Attrition through the sweeps”.

Combined, these delineate the impact of attrition on the overall makeup of the cohort for fundamental fertility and education variables. It should be noted that miscarriages have not been examined, as no comparable national statistics are available.

## METHODS

### DATA AND SOFTWARE

All available fertility-related variables from the 1958 Birth Cohort were requested from METADAC (Managing Ethico-social, Technical and Administrative issues in Data Access) and used to reconstruct in-depth fertility profiles for the 4 712 female participants in the Biomedical Sweep (Age 44) conducted in 2002. We denote by “year of Sweep” the year in which a given sweep started. Protocols for clean-up (the accuracy of which was tested using synthetic data) are supplied in the **Supplementary Material**. Variables relating to participation in sweeps, and age of leaving full-time education were also requested. A list of all 942 requested variables is provided in the **Supplementary Material**. As we were interested in the impact of attrition, we further requested information about the female cohort members who did not participate in the Biomedical Sweep (Age 44). This yielded 4 247 additional participants, with 314 variables requested (a subset of the original variable request, relating to the available sweeps). No data, if available were released for Sweep 6 (Age 42) for this group. Although we noted some slight discrepancies between the numbers of participants in our data and those recorded on published tables in Atherton et al.(2) and Power et al (1), we are in overall agreement on the productive participant counts. As part of our quality-control assessment, we noted that initial recruitment sex ratio (107.05 males to 100 females) matches the birth sex ratio of the general population. Further consistency checks are detailed in the associated R scripts.

Maternities (live births and stillbirths) and terminations as defined by the Office for National Statistics for all women living in Great Britain born in 1958 were obtained from the Office for National Statistics (ONS) and from National Records of Scotland (NRS) (35), for all years between 1972 and 2001. The ONS definitions can be found in full in ONS maternity and termination records (covering England and Wales) prior to 1992 were available only as age-group aggregates through the Conception Statistics tables (36,37). From NRS both age-group aggregates and a breakdown by mother’s age were available (35). The life table for women born in 1958 was obtained from the Human Mortality Database (38). Statistical analysis was conducted using base R (39), “tidyverse” (40), and “survival” (41) through RStudio (42). All code is available in the **Supplementary Material**.

### ANNUAL MATERNITY AND TERMINATION RATE ESTIMATES

We compared rates of maternities and terminations reported by participants with the corresponding rates in the general population, derived from official statistics. Although counts by individual years of mother’s age are available from the Scottish data, we aggregated the age groups to align them with the data releases from England and Wales. To then extract single-year estimates of maternity and termination rates for individuals born in 1958 we fitted splines, for which we tested three different specific procedures: a standard b-spline (knots at 1974, 1978, 1983, 1988, and 1993 – corresponding to the years in which participants crossed over between age classes of the vital statistics tables, for example 1974 when they entered the 16-19 years age class – a natural cubic spline (knots as in the b-spline), and a quadratic optimisation spline proposed by Grigoriev et al. (43). This last is based on minimising the sum of squared second-order differences, yielding annual rate estimates (per 1000 women) for maternity and terminations. These were compared with the per-1000-women rates for maternities and terminations within the 1958 Birth Cohort, for which we also calculated 95% confidence intervals.

### SPLINE VERIFICATION

We tested the ability of the spline methodology to recover the known individual-year counts for Scottish maternities from multi-year aggregated Scottish maternity data. We transformed the Scottish birth counts into rates per 1000 women by using the life table from the Human Mortality Database. These are plotted together with Scotland-only spline estimates in Figure 3. As the quadratic-programming spline fitted the count rates better than the natural or b-splines, we rely primarily on this method in our subsequent analysis.

**Figure 3:**
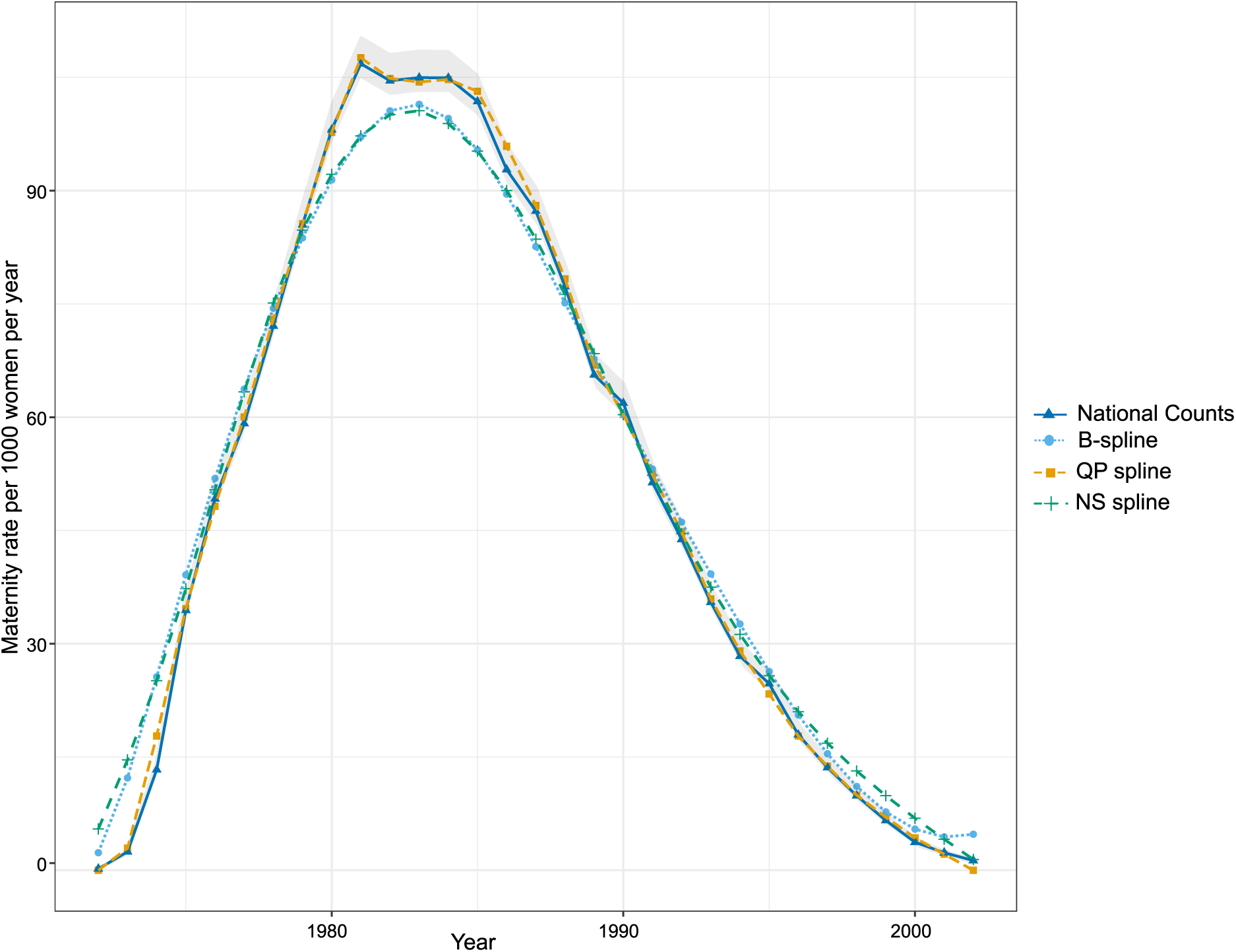
Maternity spline estimates extracted from age group aggregates against single-year counts for Scotland. Three spline methods are presented: B-splines (light blue), Quadratic programming splines (yellow), and Natural cubic splines (green). These are compared with rates per 1000 women estimated from counts (dark blue, with 95% confidence interval in light grey).

### ATTRITION THROUGH THE SWEEPS

We distinguished between two broad categories of participation: participating in the Biomedical Sweep (Age 44) (“Persisters”) or not (“Dropouts”). We define two different measures of participation: the last sweep of having participated in any way (“Overall Continuation Length”) and the last sweep of having responded to fertility-related questions (“Fertility Continuation Length”).

Although we cannot quantify possible refusal to respond to fertility questions when examining fertility-related questions, we can at least use this distinction in participation to detect patterns.

### WITHIN THE PERSISTERS

We examined associations between fertility and education event times: *age at menarche, age at first birth*, and *age left full time education*. Age at menopause was not studied as most of the sweeps occurred prior to the menopause age range. Kendall’s tau was used as a correlation measure between ages at dropout and at event. Analyses were conducted using both Continuation Length measures.

For individuals lacking event times, censoring time was determined based on auxiliary information present in the dataset. For example, for *age at first birth*, individuals with no recorded maternities were considered censored at the last time when a birth could have been observed using other fertility variables where available (such as menopause age). Where insufficient information was available to make a clear judgement about censoring time, these individuals were excluded entirely from the analysis. The numbers of excluded Persisters are summarised in Table 2.

**Table 1:** Glossary of terminology used, defining participation indicators (Persisters vs Dropouts) and measures of Continuation Length (Overall vs Fertility).

|  |  |
| --- | --- |
| PARTICIPATION INDICATORS | **Persister:** cohort member who participated in the Biomedical Sweep. |
|  | **Dropout:** cohort member who did not participate in the Biomedical Sweep. |
| MEASURES OF CONTINUATION LENGTH | **Overall Continuation Length:** last sweep in which cohort member has participated in any way |
|  | **Fertility Continuation Length:** last sweep in which cohort member has responded to any fertility-related questions |

**Table 2:** Numbers of Persisters that were included and excluded in the analyses for *age at menarche, age at first birth* and *age left full time education*.

|  | *Age at menarche* |  | *Age at first birth* |  | *Age left full time education* |  |
| --- | --- | --- | --- | --- | --- | --- |
|  | Excluded | Included | Excluded | Included | Excluded | Included |
| Overall Continuation Length | 0 | 4712 | 414 | 4298 | 239 | 4473 |
| Fertility Continuation Length | 36 | 4676 | 438 | 4274 | 275 | 4437 |

### PERSISTERS VS DROPOUTS

We compared *age at menarche* and *age at first birth* between the 4 712 Persisters and the 4 247 Dropouts using log-rank tests. *Age left full time education* was not studied here as the relevant variables were only collected in Sweep 6 (Age 42), when many Dropouts had already left the study. Age at menopause was not studied because the last records for Dropouts would be prior to age 44, which is before most women would have experienced menopause.

## RESULTS

### ANNUAL MATERNITY AND TERMINATION RATE ESTIMATES

Annual maternity rates for Persisters and Dropouts in the Birth Cohort, as well as estimates of these rates from national statistics, are shown in Figure 4. Until 1987, both Persisters and Dropouts show lower rates than national statistics, but Dropouts follow national statistics more closely than Persisters. Fewer maternities were recorded for Persisters between 1973 and 1984 than would be predicted from the records of the general population, with an average of 2.21% fewer maternities per year (sd: 0.73%, range: 1.01% - 3.13%). In 1982 and 1983 the deviation is particularly large, and shows a prominent dip in Persisters’ maternity rate curve, separating them even more from Dropouts. From 1988 onwards Persisters’ maternity rates exceed the rates calculated from national statistics.

**Figure 4:**
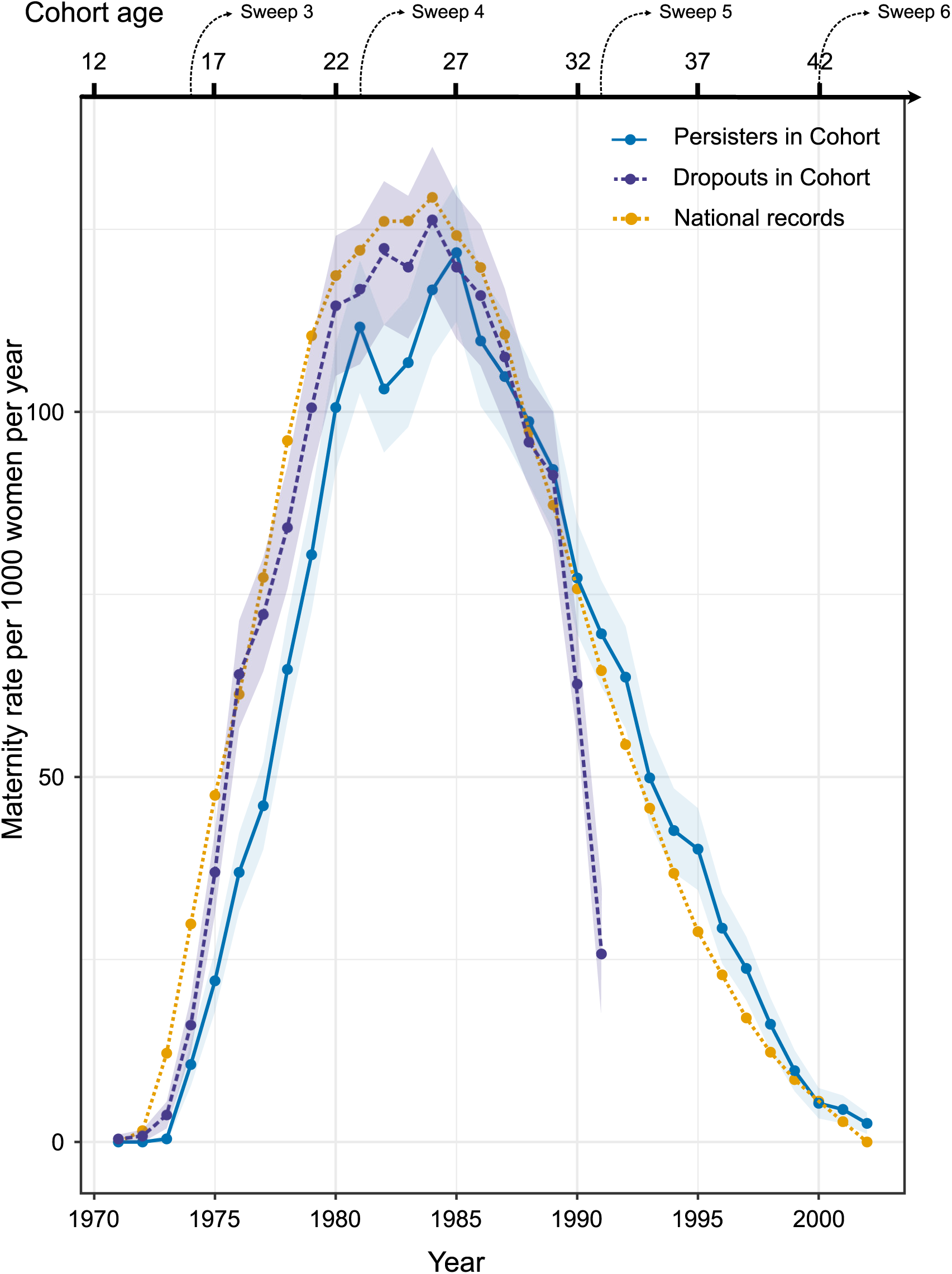
Annual maternity rates (per 1000 women) for Persisters (dark blue, with 95% confidence interval in lighter shade) and Dropouts (purple, with 95% confidence interval in lighter shade) in the Birth Cohort. These are compared to quadratic programming spline approximations of these rates based on national statistics age group aggregates (yellow).

Maternity rate differences between the cohort participants and National statistics are presented in Figure 5.

**Figure 5:**
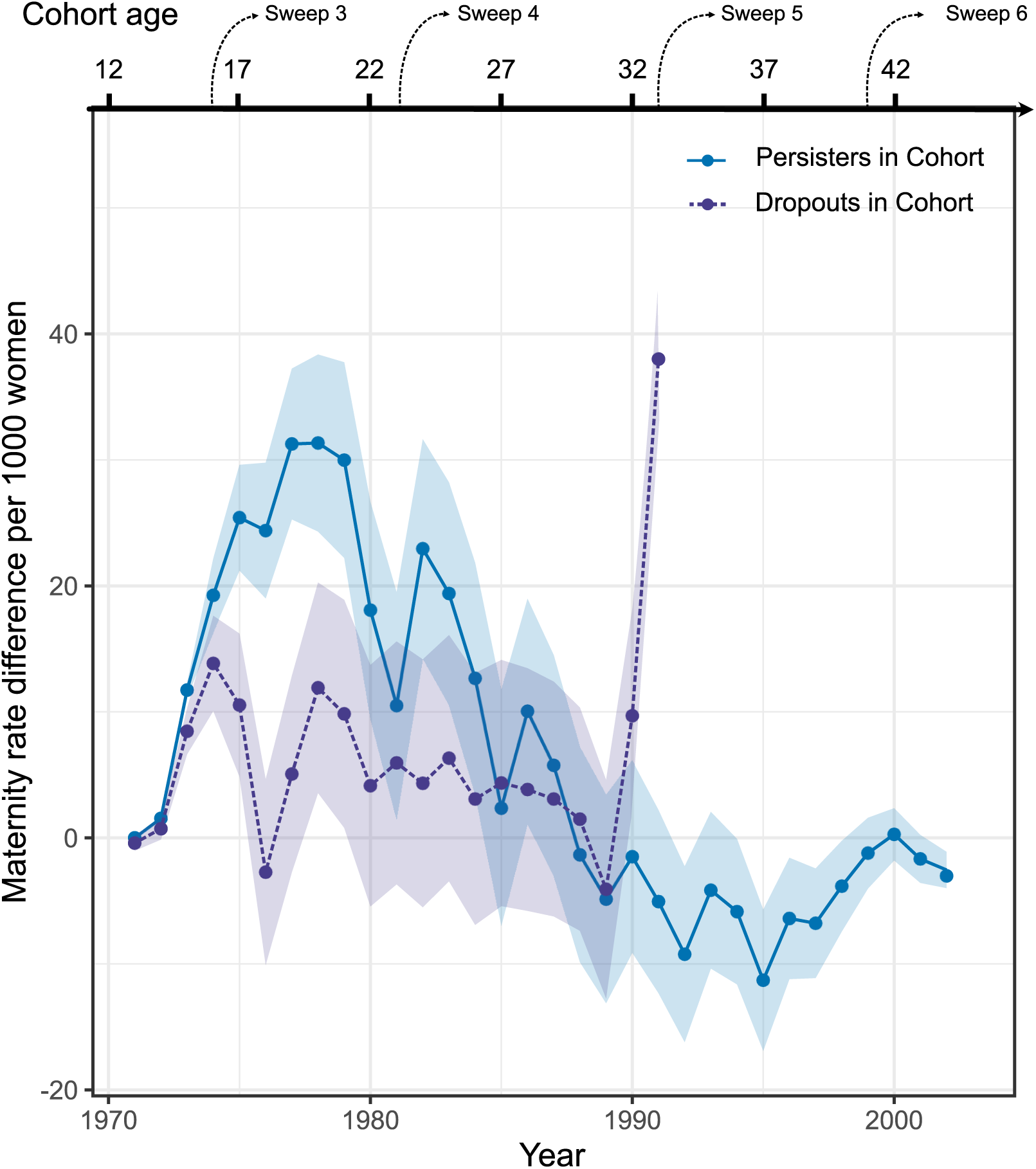
Annual maternity rate difference (per 1000 women) between National statistics and Persisters (dark blue, with 95% confidence interval in lighter shade), and National statistics and Dropouts (purple, with 95% confidence interval in lighter shade). For National statistics, rates were estimated using quadratic programming spline approximations of age group aggregates.

Comparison of termination rates between national statistics and those reported by Persisters is illustrated in Figure 6. The reported termination rates for Persisters diverge substantially from those approximated from the national statistics data. The terminations for Dropouts have not been included, as the events were not linked with specific years. This is due to the formulation of termination-related questions prior to 1990s.

**Figure 6:**
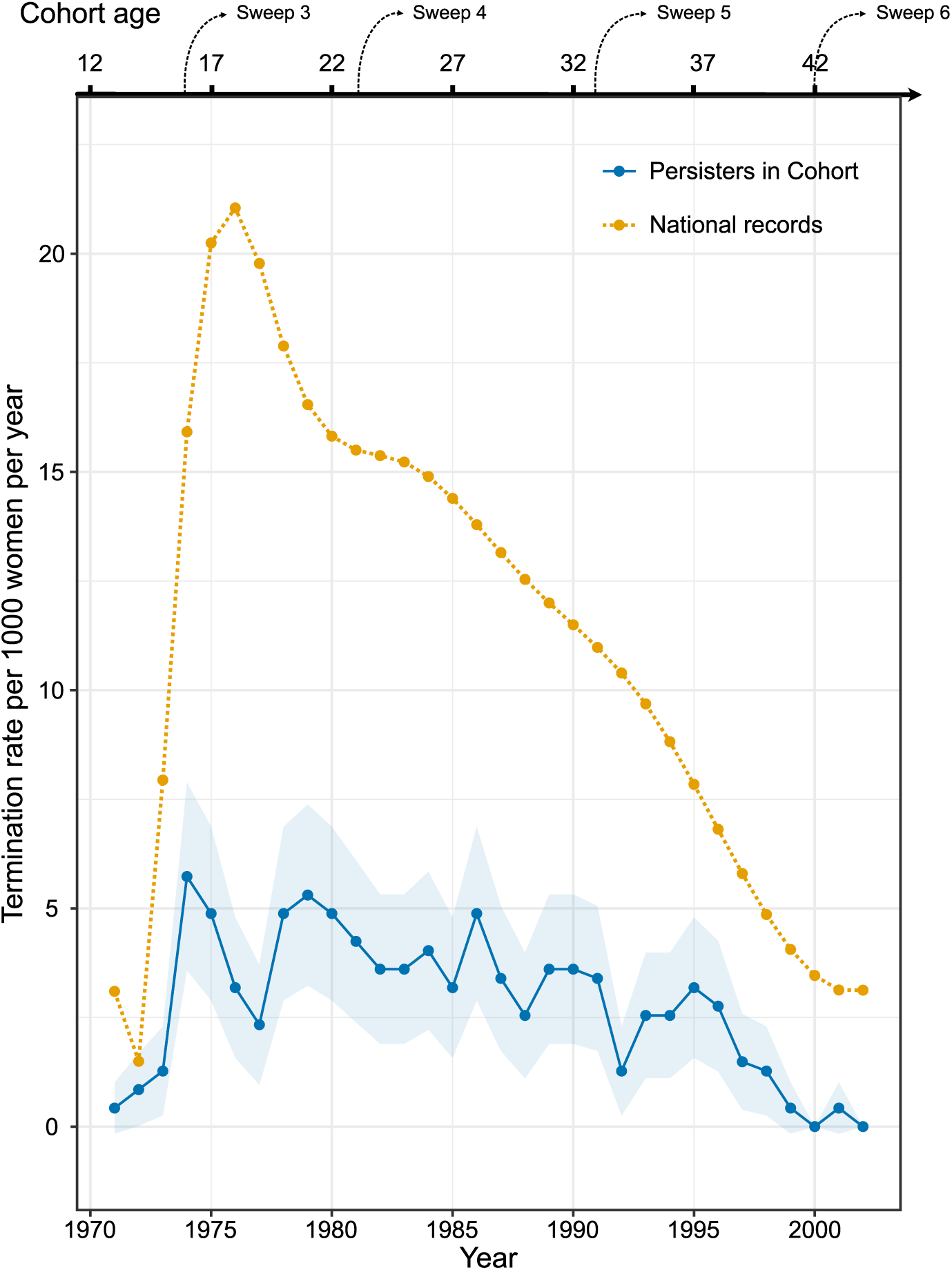
Annual termination rates (per 1000 women) as recorded for Persisters in the Birth Cohort (dark blue, with 95% confidence interval in lighter shade) compared to quadratic programming spline approximations of these rates based on national statistics age group aggregates (yellow).

### ATTRITION THROUGH THE SWEEPS

#### WITHIN PERSISTERS

Kendall’s tau between Continuation Length past the Biomedical Sweep (Age 44) and three event times – *age at menarche, age at first birth*, and *age left full time education* – are summarised in Table 3. If measured by “Overall Continuation Length”, the length of continuation evinces correlation with each of them – weakly negative for *age at menarche*, positive for *age at first birth* and *age left full time education*. In other words, members who participated in later sweeps experienced menarche at younger ages, and they both experienced their first birth and left full time education at later ages. If measured by “Fertility Continuation Length”, the negative correlation of Continuation Length with *age at menarche* disappears, but the positive correlations with the other two event times persists. This suggests that findings from the Biomedical Sweep (Age 44) on basic fertility and education characters may not be applicable to the whole cohort.

**Table 3:** Kendall’s tau rank correlation between Continuation Length and three event times: *age at menarche, age at first birth*, and *age left full time education*. τ estimates, their 95% confidence intervals, and the associated p-values were calculated for two participation indicators (Overall Continuation Length and Fertility Continuation Length) resulting in similar results for *age at first birth* and *age left full time education*, but different results for *age at menarche*.

|  | *Age at menarche* | *Age at first birth* | *Age left full time education* |
| --- | --- | --- | --- |
| Overall Continuation Length | $\tau = -0.033$<br>95% CI: (-0.045, -0.021)<br>p-value=0.009 | $\tau = 0.124$<br>95% CI: (0.112, 0.136)<br>p-value<2.2×10<sup>-16</sup> | $\tau = 0.117$<br>95% CI: (0.106, 0.127)<br>p-value<2.2×10<sup>-16</sup> |
| Fertility Continuation Length | $\tau = -0.003$<br>95% CI: (-0.02, 0.013)<br>p-value=0.778 | $\tau = 0.137$<br>95% CI: (0.12, 0.154)<br>p-value<2.2×10<sup>-16</sup> | $\tau = 0.138$<br>95% CI: (0.122, 0.153)<br>p-value<2.2×10<sup>-16</sup> |

### PERSISTERS VS DROPOUTS

Results of the comparison for *age at first birth* between Persisters and Dropouts in the Birth Cohort are presented in Figure 7. Dropouts experienced an earlier age at first birth, with median age 22, as opposed to median age 25 for Persisters.

**Figure 7:**
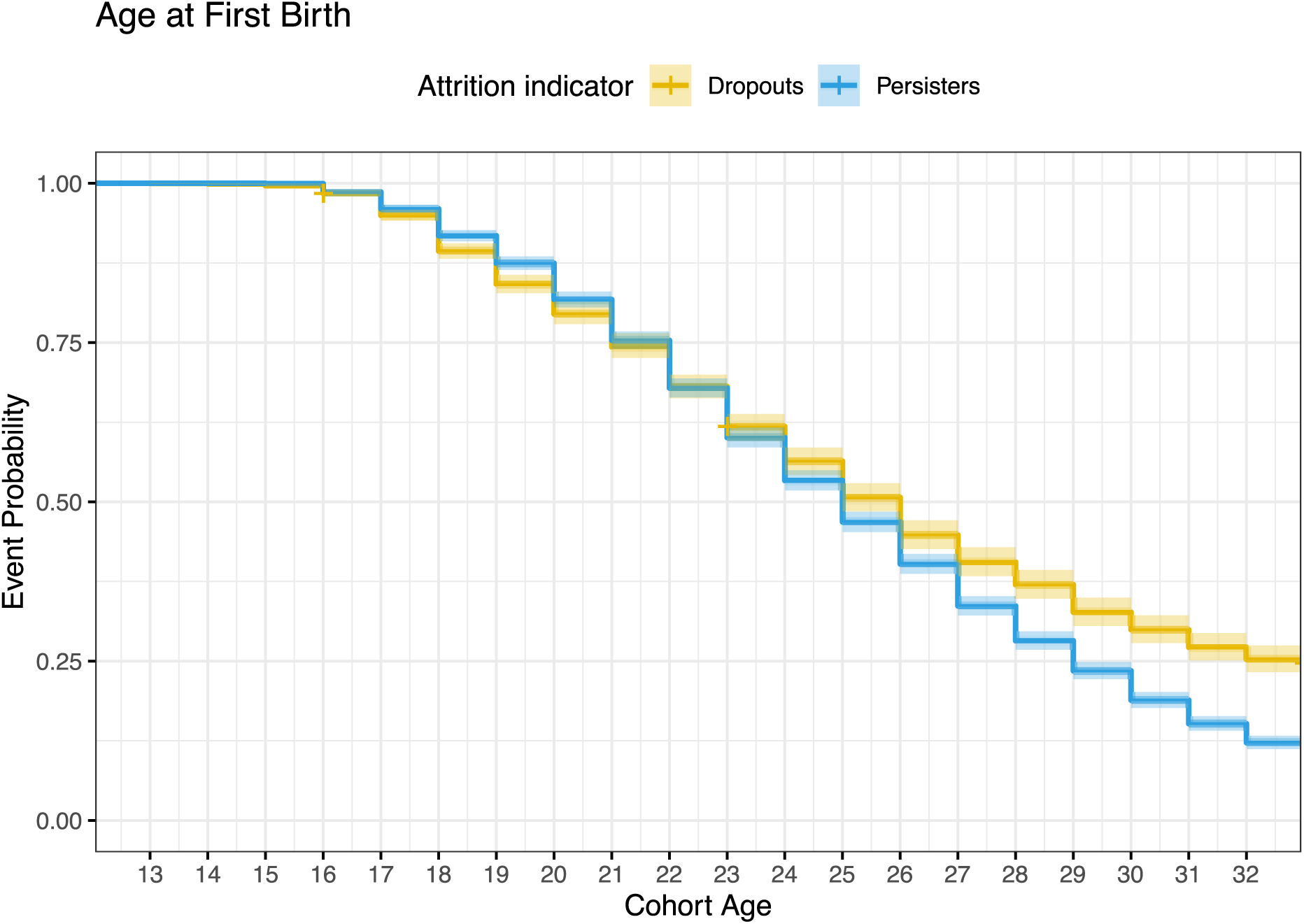
Kaplan-Meier curves for age at first birth comparison between Dropouts in the Birth Cohort (yellow) and Persisters in the Birth Cohort (in blue).

Comparison of *age at menarche* between Persisters and Dropouts in the Birth Cohort is presented in Figure 8. The two curves are similar, although proportionally more participants in the group of Persisters experience menarche at age 13.

**Figure 8:**
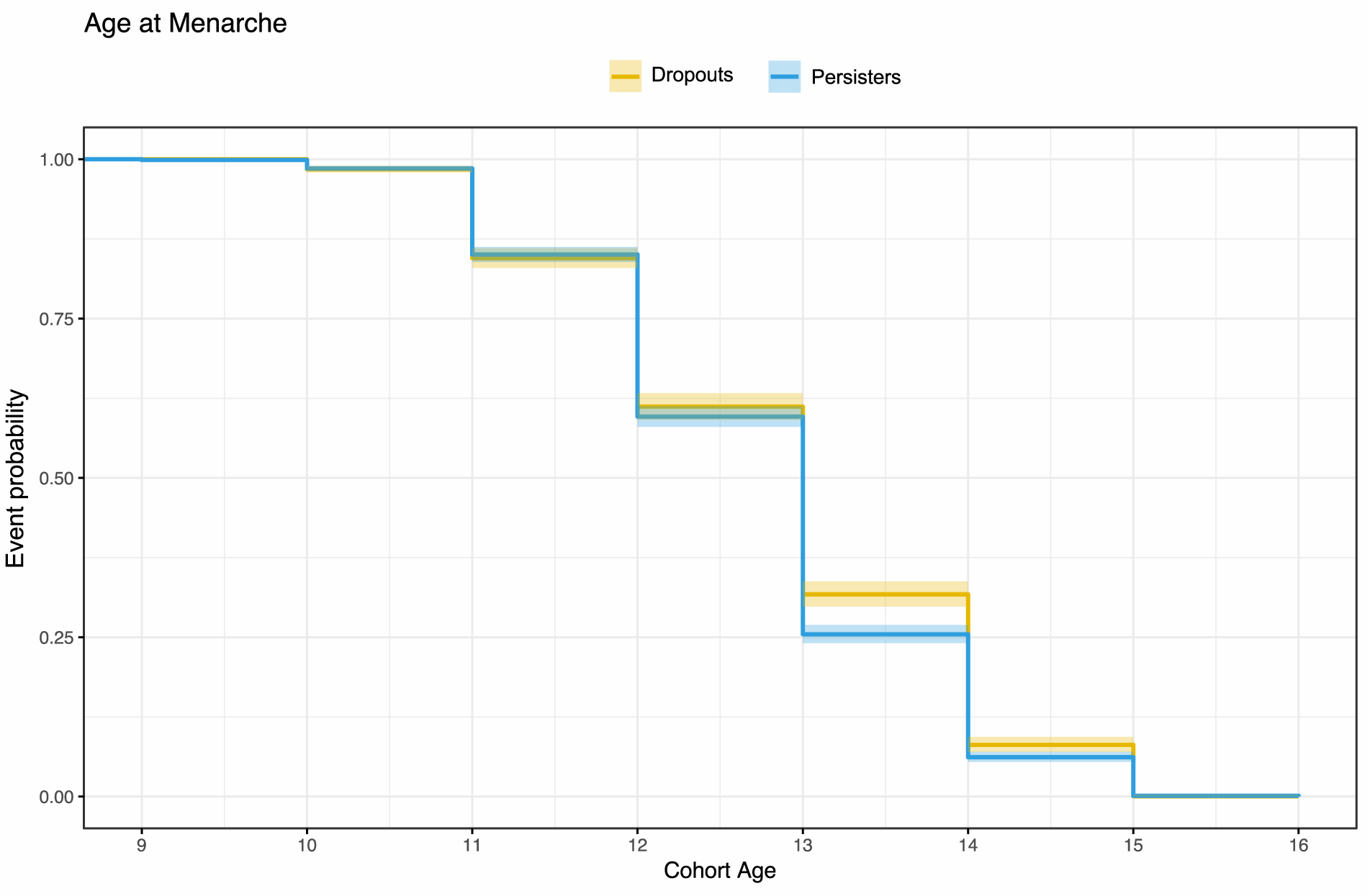
Kaplan-Meier curves for age at menarche comparison between Dropouts in the Birth Cohort (yellow) and Persisters in the Birth Cohort (blue).

## DISCUSSION

Our distinction between Persisters (women present in the Biomedical Sweep (Age 44)) and Dropouts (women who did not participate in the Biomedical Sweep (Age 44)) seems to be a productive division for understanding differential continuation rates. Suitable population estimates are often unobtainable, making direct comparisons between a study cohort and the target population impossible (44). We confirmed the utility of the quadratic-optimisation splines for recovering annual rates from events tabulated by multiyear age aggregates. We have established two key findings about the impact of attrition on basic fertility and education characteristics for the women of the 1958 Birth Cohort.

1. Fewer fertility events are reported in the cohort than would be expected from national statistics. For maternities this discrepancy lingers into the mid 1980s for Persisters and, to a lesser degree, also for Dropouts. Reported terminations for Persisters are extremely low throughout, as compared with the known rates from national statistics.
2. For Persisters both *age at first birth* and *age left full time education* are positively correlated with subjects’ length of continued study participation. Persisters experienced their first birth at a later age than Dropouts. *Age at menarche* was similar overall between the two groups.

These findings suggest that Persisters have different fertility and educational profiles to Dropouts. Furthermore, this profile diverges from national trends of their contemporaries for both maternities and terminations. Because of the nature of its initial recruitment, the 1958 Birth Cohort is often considered a representative sample of the British population. Our findings suggest that this perception is not entirely accurate, at least with regard to the fertility events considered here. Following the *Strengthening the Reporting of Observational Studies in Epidemiology (STROBE) statement* (45) we recommend that this disparity be mentioned in future work relying on the representativeness of the cohort for fertility questions.

By comparing the maternity patterns of Persisters with National Statistics we established that there were fewer maternities reported by the cohort participants between 1973 and 1984 (Figure 4). The disparity between the 1958 Birth Cohort fertility patterns and the estimates based on national statistics is perplexing. We had expected self-reported births to yield numbers and age patterns very close to those registered by statistics agencies. The discrepancy suggests either a reliability issue with the maternity records for Persisters in the Birth Cohort – underreporting or recording errors – or a genuine divergence in behaviour between Persisters and the general population. While less pronounced, Dropouts also exhibit lower maternities rates than national statistics suggest. Explanations for the observed differences need to address both the smaller numbers overall and the additional gaps due to non-random attrition. Of some interest is also the slight decrease in maternity rates for Persisters in 1982 and 1984. These are the years following Sweep 4 (Age 23), making it possible for participants to have mistakenly assumed during Sweep 5 (Age 33) that they had already reported a particular birth. This explanation does not differentiate between Dropouts and Persisters, however. As this particular dip is not observed in the Dropouts, it seems likely that there is an artefact of the data collection that affects predominantly the Persisters.

Terminations appear to be severely underreported for Persisters (Figure 6). Underreporting of terminations is potentially consequential for reproductive and medical studies (46). For example, biased termination records have led to erroneous reports of association between terminations and breast cancer, which were only later disproved through laborious large-scale studies (47,48). The underreporting is likely due to social-desirability bias. Specifically, there are two aspects of study design for the 1958 Birth Cohort that may have exacerbated it. The first is that most of the fertility data were elicited in interviews, whether face-to-face or by telephone, rather than in self-administered questionnaires which might mitigate social-desirability bias. The second is the change in fertility questionnaire structure between Sweep 4 (in 1981) and Sweep 5 (in 1991). Whereas the participants in Sweep 4 (Age 23) were asked to list any natural children they may have, in birth order, from Sweep 5 (Age 33) onwards participants were asked specifically about their pregnancies, starting from the most recent and going backward in time. Miscarriages and terminations from Sweep 4 (Age 23) are reported only collectively, as total numbers, without associated dates. National statistics (Figure 6) suggest that the terminations for women born in 1958 peaked by 1976. This means that the increased resolution in data collection for the cohort, introduced in Sweep 5 (Age 33), appears too late. The responses to the pregnancy questions from 1991 onwards, although informative in assisting to reconstruct the past, may be compromised by recall bias on top of the inevitable social-desirability bias, and the aforementioned non-random attrition.

For Persisters, we have found that participation in the final four sweeps (Biomedical (Age 44) and Sweeps 7-9 (Ages 46-55)) was associated with being older both at first birth and at departure from full-time education. That these two event times follow similar patterns might have been anticipated from positive correlation established in the literature (49,50), but we know of no reason to have predicted an association between the timings of those events and Continuation Length in the study.

The correlation between Continuation Length and the above-mentioned two event times becomes stronger when continuation is defined by a subject’s engagement specifically with the fertility questions. How to measure Continuation Length is an important consideration, especially within the context of any statistical analyses of fertility – for instance, comparing menopause timing – that try to account for right censoring of missing subjects. Using Overall Continuation Length as the censoring time for individuals who lack event times may be misleading if the individuals have not interacted with any of the fertility questions. Fundamentally, the last time when we can confidently assume that an event has not occurred is the last time when we have been explicitly or implicitly told it has not occurred. When subjects have responded only to some questions we may be left unsure whether they are, in fact, censored. As in our case menopause timings are not captured by an external registry, the suggested correction proposed by Lesko et al. (51) does not apply.

Whether overall the 1958 British Birth Cohort is a good proxy for the British population of this age group – in other words, whether it is representative of the population – is a multifaceted question that defies any simple answer. The general definition of representativeness of cohorts in research on human populations, and whether or not it is an essential quality of a study, has been discussed extensively (25–30). The analyses presented here illustrate some of the ways in which representativeness may depend on the research question under consideration. This has implications for the use of the cohort study data. Once a research question has been defined, relevant biases can be established and, in some cases, quantified, for example by means of causal diagrams, relative odds ratios, or expert assessment (52,53). Then, depending on the combined nature of the bias and the research question, remedial action can be considered – whether it is a quantitative correction such as multiple imputation with inverse probability weighting (54,55), or by other means, such as those suggested in (56). Since it is *a priori* not known whether the effect of biases in associations are very modest, as in (52), or quite substantial, as in (54), it is important to determine how each specific research question may interact with the identified biases.

## Data Availability

Data governance was provided by the METADAC data access committee, funded by ESRC, Wellcome, and MRC. (Grant Number: MR/N01104X/1)
This work made use of data and samples generated by the 1958 Birth Cohort (NCDS), which is managed by the Centre for Longitudinal Studies at the UCL Institute of Education, funded by the Economic and Social Research Council (grant number ES/M001660/1). Access to these resources was enabled via the Wellcome Trust & MRC: 58FORWARDS grant [108439/Z/15/Z] (The 1958 Birth Cohort: Fostering new Opportunities for Research via Wider Access to Reliable Data and Samples). Before 2015 biomedical resources were maintained under the Wellcome Trust and Medical Research Council 58READIE Project (grant numbers WT095219MA and G1001799).

## ETHICS

Access to anonymised linked records from NCDS was obtained after approval by the METADAC panel on April 11th 2017 (application number: MDAC-2017-0007-01).

## ACKNOWLEDGMENTS

Data governance was provided by the METADAC data access committee, funded by ESRC, Wellcome, and MRC. (Grant Number: MR/N01104X/1)

This work made use of data and samples generated by the 1958 Birth Cohort (NCDS), which is managed by the Centre for Longitudinal Studies at the UCL Institute of Education, funded by the Economic and Social Research Council (grant number ES/M001660/1). Access to these resources was enabled via the Wellcome Trust & MRC: 58FORWARDS grant [108439/Z/15/Z] (The 1958 Birth Cohort: Fostering new Opportunities for Research via Wider Access to Reliable Data and Samples). Before 2015 biomedical resources were maintained under the Wellcome Trust and Medical Research Council 58READIE Project (grant numbers WT095219MA and G1001799).

